# Association of Cardiologist Clinic Visits with Cardiovascular Primary Prevention Outcomes Among People with HIV from Underrepresented Racial and Ethnic Groups in the Southern United States

**DOI:** 10.1101/2024.08.08.24311709

**Authors:** Matthew S. Durstenfeld, C. Larry Hill, Robert M. Clare, Karen Chiswell, Gretchen Sanders, Shamea Gray, Linda Cooper, Joseph Vicini, Keith Marsolo, Nwora Lance Okeke, Eric G. Meissner, Kevin L. Thomas, Caryn G. Morse, Gerald S. Bloomfield, April C. Pettit, Chris T. Longenecker

**Author notes:** **Address for Correspondence:** Matthew S. Durstenfeld, MD MAS, Division of Cardiology, UCSF at Zuckerberg San Francisco General Hospital 1001 Potrero Avenue, 5G8 San Francisco, CA 94110, USA.

## Abstract

**Background:** People with HIV (PWH) are at elevated risk for atherosclerotic cardiovascular disease (ASCVD). Underrepresented racial and ethnic groups (UREGs) with HIV in the southern U.S. are disproportionately affected, yet whether cardiology specialist care for this at-risk group improves blood pressure and lipid control or prevents cardiovascular events is unknown.

**Methods:** We evaluated a cohort of PWH from UREGs at elevated ASCVD risk without known cardiovascular disease who received HIV-related care from 2015–2018 at four academic medical centers in the Southern United States with follow up through 2020. Primary outcomes were blood pressure control (<140/90 mmHg) and lipid control (LDL-C ≤ 100 mg/dl) over 2 years and time to first major adverse cardiovascular (MACE) event. Statistical analyses were adjusted for cohort/site and patient factors including HIV measures and comorbidities.

**Results:** Among 3972 included PWH (median age 47 years old, 32.6% female) without diagnosed cardiovascular disease, 276 (6.9%) had a cardiology clinic visit. Cardiology clinic visits were not significantly associated with subsequent blood pressure control (adjusted OR 0.78, 95% CI 0.49-1.24, p=0.29) or lipid control (adjusted OR 2.25, 95% CI 0.72-7.01, p=0.16). Over a median follow up of 5 years, patients who had a cardiology clinic visit had higher risk of MACE, overall mortality, and falsification endpoints (hospitalization or death from accident/trauma and pneumonia/sepsis) indicating a higher risk group overall, even after adjusting for measured risk factors.

**Conclusions:** Among UREG PWH at elevated cardiovascular risk, a cardiology clinic visit was not associated with improved cardiovascular risk factors or reduced risk of cardiovascular events. Our study suggests that seeing a cardiologist is not alone sufficient to promote cardiovascular health or prevent cardiovascular events among PWH, but with low confidence given the higher risk among those who had a cardiology visit.

**What is known?:**

- People with HIV are at increased cardiovascular risk, and the burden of both cardiovascular disease and HIV are high among people from underrepresented racial and ethnic groups who live in the Southern United States.
- Treating people with HIV at elevated cardiovascular risk with statins reduces risk of cardiovascular events.

**What the study adds?:**

- Among people with HIV at elevated cardiovascular risk from underrepresented racial and ethnic groups who received care at four academic medical centers in the southern United States, cardiology clinic visits were not associated with better lipid control, blood pressure control, or prevention of cardiovascular events.
- People with HIV who attended a cardiology clinic visit had higher risk of cardiovascular events and mortality.

## Introduction

People with HIV (PWH) have elevated risk of developing atherosclerotic cardiovascular disease (ASCVD), with two-fold higher risk of myocardial infarction.^1^ Similarly, PWH are at increased risk for heart failure and arrhythmias. In the United States, people who belong to underrepresented racial and ethnic groups (UREGs) bear a disproportionate share of both HIV and ASCVD disease burden, especially in the South.

Among PWH, early diagnosis of HIV, linkage to HIV care, and treatment with antiretroviral therapy (ART) remain foundational to promote long and healthy lives. Among PWH linked to HIV care, guidelines recommend estimating ASCVD risk to guide primary prevention strategies.^2,3^ Among those predicted to have elevated risk, one potential strategy to mitigate the excess risk of ASCVD may be involvement of cardiologists in the care of PWH to optimize primary prevention of cardiac events. Whether a clinic visit with a cardiologist has an impact on primary prevention of cardiovascular outcomes among PWH (or even the general population without HIV) is unknown.

Therefore, we designed a multi-center observational cohort study to examine the association between cardiology clinic visits and cardiovascular outcomes among PWH from UREGs with elevated ASCVD risk. We hypothesized that a clinic visit with a cardiologist would be associated with improvement in risk factors (blood pressure and lipid) control at 2 years and therefore lower risk of major adverse cardiovascular events and mortality over 5 years among PWH from UREGs. Because our research question is causal (“Does seeing a cardiologist prevent heart disease?”), we employed multiple comparative effectiveness strategies to rigorously answer these questions, as described further in the methods section below. Our hope was that our findings would inform whether a cardiologist visit should be implemented as a strategy to prevent cardiovascular events among PWH from UREGs.

## Methods

### Study Design

As previously described, the Pathways to Cardiovascular Disease Prevention and Impact of Specialty Referral in Under-Represented Racial/Ethnic Minorities with HIV (PATHWAYS) study (NCT04025125) is a multi-center collaborative observational electronic health record (EHR) based cohort study focused on PWH who are UREGs who receive clinical care in the Southern United States.^4^ For this study, we used a series of nested cohort studies, as described in more detail in the Statistical Analysis section below.

### Setting

Four academic health centers located in the United States South that participate in the Stakeholders, Technology, and Research (STAR) Clinical Research Network were included in this study: Duke Health, The Medical University of South Carolina, Vanderbilt University Medical Center, and Wake Forest Baptist Health. EHR data was harmonized in the National Patient-Centered Clinical Research Network, PCORnet® Common Data Model.^5^ We included EHR data from January 1, 2014-December 31, 2020 extracted and harmonized from 2021-2022.

### Participants

We included individuals aged 18-99 years old with evidence of HIV documented in the EHR who were retained in HIV care at a participating site (defined by an HIV viral load laboratory test, a prescription for antiretroviral therapy (ART), and/or an encounter with an HIV provider in the 12 months prior to their index date) and with race or ethnicity documented as Black/African American, American Indian or Alaska Native, Asian, Multiple Race, or Hispanic. We included those who had elevated ASCVD risk defined as a 10-year risk ≥5% estimated by the pooled cohort equations or ≥7.5% by the Framingham risk score for those ≥40 years of age, and ≥40% lifetime risk by the pooled cohort equations for those < 40 years of age. The index date for entry into the overall cohort was defined as the first clinic visit between 2015-2018 with elevated ASCVD risk. We excluded those with prior major adverse cardiac event (MACE) including prior myocardial infarction or stroke (requiring secondary prevention), heart failure, and atrial fibrillation, or any prior encounter with a cardiologist within one of the participating centers with a lookback period to 2014. We also excluded those who were documented to have MACE, heart failure, or atrial fibrillation at the time of their cardiologist visit. The CONSORT Diagram is shown in Supplemental Figure 1. Patients were followed for up to five years, until death, or until they were administratively censored at the end of EHR data availability (December 31, 2020).

### Exposure, Outcomes, and Other Variable Definitions

The exposure was an ambulatory clinic visit with a cardiologist, as defined previously based on the provider specialty listed in the common data model and the provider’s National Provider Identifier restricted to ambulatory clinic visits and excluding encounters for cardiac diagnostic testing alone,^4^ that occurred after the index date and before the end of follow up. The comparator group consisted of patients with at least one ambulatory clinic visit after the index date and before the end of follow up with at least one non-cardiologist clinician. The selection of patients into the exposure and comparator groups is described under Statistical Methods. For the MACE outcome, we further stratified the exposure into cardiology visits for “prevention” and “management” based on the ICD codes documented at the cardiology visit. All ICD codes coded for during a cardiology visit were manually classified by two board-certified cardiologists blinded to clinical data with disagreements resolved through consensus (Supplemental Materials Appendix 1-3). We classified visits which had ICD codes for cardiovascular diagnoses, abnormal cardiovascular testing, and cardiovascular symptoms (chest pain, for example), as management visits. We classified visits which only had ICD codes for non-cardiovascular diagnoses, cardiovascular risk factors, and pre-operative assessment as prevention visits.

Our first two primary outcomes were blood pressure control (defined as systolic blood pressure <140 mm Hg and diastolic blood pressure <90 mm Hg) and lipid control defined as LDL cholesterol ≤ 100 mg/dl (2.6 mmol/L). For blood pressure we averaged available blood pressures within each given calendar month and used the last available month within two years of the cardiology visit or equivalent non-cardiology visit to define control. For lipids we used the last available lipid panel after the cardiology visit or equivalent non-cardiology visit within two years.

The third primary outcome was incidence of major adverse cardiovascular events (MACE) over five years defined as cardiovascular death, myocardial infarction, acute coronary syndrome, stroke including transient ischemic attack, peripheral arterial disease treated with revascularization with percutaneous or surgical bypass, and coronary artery disease treated with revascularization with percutaneous coronary intervention or coronary artery bypass grafting. Non-fatal outcomes were defined by hospitalization discharge ICD codes for diagnoses and CPT codes for procedures and were not manually adjudicated due to lack of access to clinical notes. Cardiovascular death was defined based on ICD codes for cardiovascular causes of death recorded in the National Death Index Plus.

As secondary outcomes we also considered the change in LDL and change in systolic blood pressure over 2 years as continuous measures assessed longitudinally among those with two years of follow-up available. We also considered heart failure hospitalization, all-cause mortality, and alternative definitions of MACE including: MACE + heart failure hospitalization and MACE + non-cardiovascular mortality.

We included comorbid conditions and procedures recorded in the electronic health record as covariates using ICD codes and the Charlson Comorbidity Index (Supplemental Table 1). We also included medication prescriptions for antiretroviral therapy and cardiovascular medications, systolic and diastolic blood pressures, body mass index, and laboratory results including lipid, creatinine (to estimate the glomerular filtration rate based on the 2009 version of the CKD EPI equation without race), and hemoglobin A1c.

### Data Sources

Data were extracted from the EHRs of the participating sites and harmonized into a single dataset. Data included participant level data (demographics) as well as billing code (ICD/CPT) data, vital signs, and laboratory data. Mortality data, including cause of death, were obtained from the National Death Index Plus.

### Statistical Methods

#### Sequence of nested cohort studies

To select the cardiology group and a non-cardiology comparator group, we divided the study period (2015-2018) into eight six-month intervals. In each interval we constructed a cohort study (Figure 1). For each cohort we identified living participants who met the eligibility criteria in that period and had not yet had a MACE event, or a prior cardiology visit. We considered those “exposed” if they had a clinic visit with a cardiologist during that period using the first cardiology visit date as the start of follow-up time. We classified those without a cardiology encounter during that period as “unexposed” and started their follow-up time at the last outpatient (non-cardiology) visit date in that six-month period. Patients who were “unexposed” were eligible for inclusion in the subsequent cohort if they continued to meet eligibility criteria, thus patients could be counted in multiple cohorts. We then pooled the eight sequential cohort studies into a single analysis, accounting for inclusion of the same patient in multiple cohorts.^6^ This pooled cohorts design allowed us to ensure that both exposed and unexposed patients were still eligible at the time of entry into the cohort; provided a mechanism to align the start of follow-up between exposed and unexposed groups; and minimized immortal time bias that could result from assigning patients to groups after the start of follow-up (for example, if follow-up had been started prior to cardiology visit at the time when ASCVD risk was elevated).

**Figure 1.**
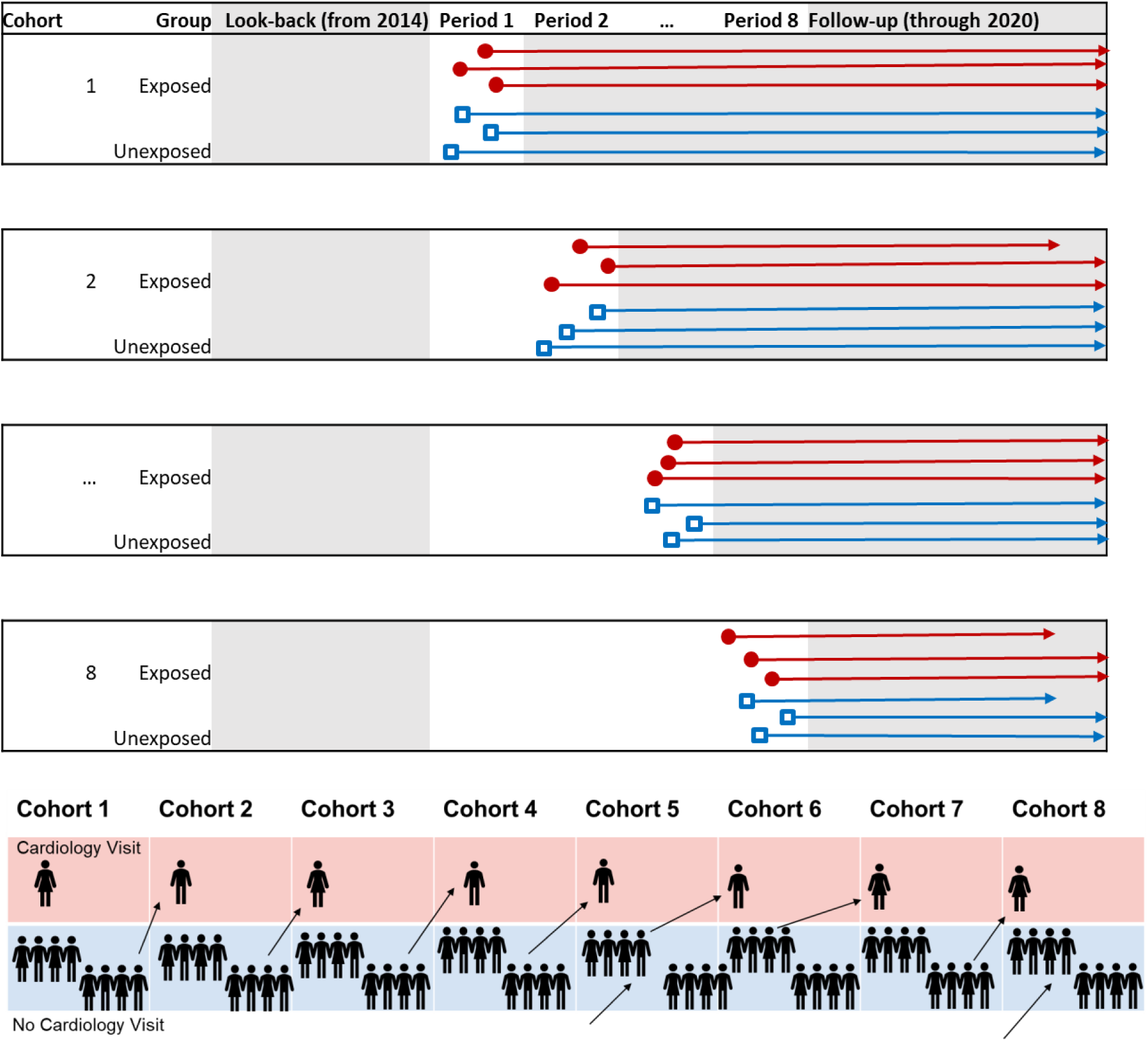
**Diagram of Study Participants** We used a sequential cohort design in which the study period was divided into eight six-month intervals. Within each interval we constructed a cohort study of eligible patients, identifying patients with a cardiology visit (exposed group), with remaining patients comprising the unexposed group, excluding those with MACE or a cardiology visit during the lookback period. For each cohort, follow-up started at the time of the first cardiology visit in the interval (red circle), or the last non-cardiology visit in the interval (blue square), thus minimizing the immortal time from the delay between eligibility and first cardiology encounter. As shown in the bottom panel, patients with a cardiology clinic visit (exposed) were ineligible for subsequent cohorts, but patients without a cardiology visit (unexposed) could be included in the subsequent cohort if they continued to meet eligibility criteria (e.g., primary prevention status).

#### Baseline Data

Baseline data were assessed for each of the eight cohorts. Vital signs, LDL, and cardiovascular medications were assessed at/before the start of follow-up within each cohort (up to 2 years prior for LDL and blood pressure and up to 13 months prior for medications). Other characteristics were assessed using the most recent available data up to the end date of the cohort for which that patient was included.

#### Missing Data

For missing data on covariates and primary prevention status (no history of MACE), we carried forward the last available data based on the baseline date for each sub-cohort. Because we used mixed effects models for the longitudinal data, we did not impute missing outcomes (LDL cholesterol, for example).

#### Models

To estimate associations between cardiology encounters and subsequent lipid and blood pressure changes, we used hierarchical mixed effects models with random effects for patients, and patients nested within a cohort to account for the potential inclusion of a single patient in multiple cohorts as well as longitudinal measures for a patient within a cohort. For time-to-first-MACE analyses, we used hierarchical Cox proportional hazards models using a three-level exposure variable (cardiovascular prevention encounter, cardiology management encounter, and no cardiology encounter) based on the first recorded cardiology encounter, censored participants at the time of first MACE, and used robust sandwich covariance estimates to account for potential contribution of the same participant across cohorts. Adjustment variables included site of care, age, sex, insurance, rural location, social deprivation index, HIV variables (CD4 count, viral suppression, anti-retroviral therapy), hepatitis C, Charlson Comorbidity Index, estimated glomerular filtration rate, BMI, diabetes, and smoking. For the lipid analyses we additionally adjusted for baseline LDL-C and lipid lowering medication use and the blood pressure analyses for baseline systolic blood pressure and anti-hypertensives. For MACE we additionally adjusted for SBP, LDL-C, antihypertensive treatment, and lipid therapy.

#### Falsification Endpoints

We hypothesized that cardiology encounters would be unlikely to have an impact on HIV viral suppression (defined as viral load <200 copies/ml) or change in CD4 counts. We also hypothesized that cardiology encounters would be unlikely to impact hospitalization or death from pneumonia or sepsis or from accident, suicide, or homicide. Therefore, we used falsification endpoints as negative controls to detect confounding by being unaffected by the exposure of interest but likely reflecting an intrinsic quality that affects the outcomes of interest.^7^

### Study Approval and Reporting

The Duke University Health System IRB approved this study with a waiver of informed consent as the single IRB with the other sites relying on this IRB. The study results are reported in accordance with the Strengthening the Reporting of Observational Research guidelines.^8^

## Results

We identified 3972 individuals who met our inclusion and exclusion criteria including 276 (6.9%) who had a cardiology clinic visit (Table 1). Approximately one third of the cohort were female sex, nearly all identified as Black (95%), and 9% identified their ethnicity as Hispanic or Latinx. There were notable differences between those who did and did not have a cardiology clinic visit. For example, those with a cardiology clinic visit were more likely to be older, female, and have diabetes or have a history of an AIDS diagnosis, suggestive of more advanced HIV disease (lower nadir CD4 count and/or opportunistic infection or malignancy). They were also more like to have a higher predicted ASCVD risk by the pooled cohort equations (median 9.0% over 10 years compared to median 6.6%) and higher median Charlson comorbidity index (5 versus 3). There were no differences in the proportion prescribed antiretroviral therapy, CD4 count, or the proportion virally suppressed.

**Table 1:** Participant Characteristics by Cardiology Encounter.

| Characteristic<sup>[1]</sup> | No Cardiology Visit<br>N=3696 | ANY Cardiology<br>Visit<br>N=276<sup>[5]</sup> | Cardiology-Preventive<br>N=154<sup>[5]</sup> | Cardiology-Management<br>N=122<sup>[5]</sup> |
| --- | --- | --- | --- | --- |
| Site, n (%) |  |  |  |  |
| Duke | 924 (25.0) | 51 (18.5) | 25 (16.2) | 26 (21.3) |
| MUSC | 700 (18.9) | 22 (8.0) | 11 (7.1) | 11 (9.0) |
| Vanderbilt | 1208 (32.7) | 47 (17.0) | 7 (4.5) | 40 (32.8) |
| Wake Forest | 864 (23.4) | 156 (56.5) | 111 (72.1) | 45 (36.9) |
| **Demographics** |  |  |  |  |
| Age, years, Median (IQR) | 47.0 (36.0, 54.0) | 53.0 (46.0, 58.0) | 52.5 (46.0, 57.0) | 53.0 (46.0, 58.0) |
| Race, n/N (%) |  |  |  |  |
| American Indian or Alaska Native | 16/3440 (0.5) | 2/259 (0.8) | 2/145 (1.4) | 0/114 (0.0) |
| Asian | 55/3440 (1.6) | 2/259 (0.8) | 0/145 (0.0) | 2/114 (1.8) |
| Black or African American | 3257/3440 (94.7) | 248/259 (95.8) | 142/145 (97.9) | 106/114 (93.0) |
| Native Hawaiian or Other Pacific Islander | 2/3440 (0.1) | 0/259 (0.0) | 0/145 (0.0) | 0/114 (0.0) |
| White | 94/3440 (2.7) | 7/259 (2.7) | 1/145 (0.7) | 6/114 (5.3) |
| Multiple race | 16/3440 (0.5) | 0/259 (0.0) | 0/145 (0.0) | 0/114 (0.0) |
| Hispanic or Latino/Latina/Latinx Ethnicity, n/N (%) | 322/3687 (8.7) | 21/275 (7.6) | 10/153 (6.5) | 11/122 (9.0) |
| Sex, n/N (%) |  |  |  |  |
| Female | 1174/3696 (31.8) | 119/276 (43.1) | 74/154 (48.1) | 45/122 (36.9) |
| Male | 2522/3696 (68.2) | 157/276 (56.9) | 80/154 (51.9) | 77/122 (63.1) |
| Urban area (vs Rural), n (%) | 3276 (88.7) | 263 (95.3) | 145 (94.2) | 118 (96.7) |
| Social Deprivation Index | 76.0 (56.0, 89.0) [3616] | 76.0 (57.0, 93.0) [268] | 78.5 (60.0, 94.0) [148] | 74.0 (51.5, 92.0) [120] |
| **Insurance** |  |  |  |  |
| Medicare/Medicaid, n/N (%) | 1550/3696 (41.9) | 168/276 (60.9) | 110/154 (71.4) | 58/122 (47.5) |
| Private insurance, n/N (%) | 1575/3696 (42.6) | 98/276 (35.5) | 40/154 (26.0) | 58/122 (47.5) |
| HRSA insurance, n/N (%) | 990/3696 (26.8) | 46/276 (16.7) | 27/154 (17.5) | 19/122 (15.6) |
| Other insurance, n/N (%) | 179/3696 (4.8) | 10/276 (3.6) | 5/154 (3.2) | 5/122 (4.1) |
| None or missing insurance, n/N (%) | 1986/3696 (53.7) | 197/276 (71.4) | 125/154 (81.2) | 72/122 (59.0) |
| **Vital Measures** |  |  |  |  |
| BMI, kg/m<sup>2</sup>, Median (IQR) [N] | 27.4 (23.7, 32.3) [3326] | 27.5 (23.3, 33.6) [273] | 27.3 (22.7, 33.7) [152] | 27.6 (24.5, 33.5) [121] |
| **Medication Prescription prior to 'Baseline' Visit<sup>[2]</sup>** |  |  |  |  |
| Diabetes medication, n (%) | 220 (6.8) | 26 (10.5) | 15 (10.9) | 11 (10.1) |
| Antihypertensive, n (%) | 902 (27.9) | 102 (41.3) | 56 (40.6) | 46 (42.2) |
| Lipid Lowering, n (%) | 396 (12.2) | 53 (21.5) | 26 (18.8) | 27 (24.8) |
| Anticoagulation, n (%) | 106 (3.3) | 6 (2.4) | 2 (1.4) | 4 (3.7) |
| Antiplatelet, n (%) | 137 (4.2) | 22 (8.9) | 10 (7.2) | 12 (11.0) |
| **Medication Prescription on/after 'Baseline' Visit<sup>[2]</sup>** |  |  |  |  |
| Diabetes medication, n (%) | 253 (7.8) | 30 (12.1) | 15 (10.9) | 15 (13.8) |

| **Characteristic<sup>[1]</sup>** | **No Cardiology Visit**<br>**N=3696** | **ANY Cardiology**<br>**Visit**<br>**N=276<sup>[5]</sup>** | **Cardiology-**<br>**Preventive**<br>**N=154<sup>[5]</sup>** | **Cardiology-**<br>**Management**<br>**N=122<sup>[5]</sup>** |
| --- | --- | --- | --- | --- |
| Antihypertensive, n (%) | 926 (28.6) | 126 (51.0) | 60 (43.5) | 66 (60.6) |
| Lipid Lowering, n (%) | 481 (14.9) | 59 (23.9) | 25 (18.1) | 34 (31.2) |
| Anticoagulation, n (%) | 103 (3.2) | 15 (6.1) | 9 (6.5) | 6 (5.5) |
| Antiplatelet, n (%) | 140 (4.3) | 29 (11.7) | 10 (7.2) | 19 (17.4) |
| **Other Relevant Medical History** |  |  |  |  |
| Hypertension, n (%) | 1425 (38.6) | 185 (67.0) | 109 (70.8) | 76 (62.3) |
| Diabetes, n (%) | 483 (13.1) | 67 (24.3) | 38 (24.7) | 29 (23.8) |
| Total Cholesterol > 200 mg/dL, n (%) | 785 (27.2) | 68 (27.0) | 36 (25.5) | 32 (28.8) |
| Hepatitis C, n (%) | 419 (11.3) | 41 (14.9) | 23 (14.9) | 18 (14.8) |
| Charlson Comorbidity Index (Glasheen 2019), score [N]<sup>[3]</sup> | 3.0 (3.0, 6.0) [3696] | 5.0 (3.0, 7.0) [276] | 6.0 (3.0, 8.0) [154] | 5.0 (3.0, 7.0) [122] |
| Estimated Glomerular Filtration Rate, mL/min/1.73m<sup>2</sup>, Median (IQR) [N]<sup>[4]</sup> | 86.5 (69.6, 101.4) [3559] | 78.8 (64.5, 98.5) [273] | 79.1 (65.8, 101.9) [153] | 75.6 (63.4, 93.2) [120] |
| **HIV Characteristics** |  |  |  |  |
| AIDS diagnosis, n (%) | 988 (26.7) | 107 (38.8) | 67 (43.5) | 40 (32.8) |
| CD4 Count, cells/uL Median (IQR) [N] | 598.0 (380.0, 829.0) [3373] | 610.5 (370.0, 860.0) [256] | 560.0 (332.0, 820.0) [141] | 670.0 (395.0, 896.0) [115] |
| CD4 count <200 in previous 2 years, n/N (%) | 566/3373 (16.8) | 45/256 (17.6) | 30/141 (21.3) | 15/115 (13.0) |
| HIV viral load, copies/mL, Median (IQR) [N] | 40.0 (2.3, 100.0) [2537] | 24.0 (1.6, 97.7) [211] | 20.0 (1.0, 251.2) [123] | 39.4 (4.1, 54.8) [88] |
| **Viral Suppression, n (%)** |  |  |  |  |
| Undetectable | 1159 (31.4) | 65 (23.6) | 31 (20.1) | 34 (27.9) |
| <200 copies/mL | 2018 (54.6) | 167 (60.5) | 91 (59.1) | 76 (62.3) |
| >=200 copies/mL | 519 (14.0) | 44 (15.9) | 32 (20.8) | 12 (9.8) |
| Antiretroviral Therapy, n (%) | 2986 (80.8) | 229 (83.0) | 126 (81.8) | 103 (84.4) |
| **Behavioral and Social Factors** |  |  |  |  |
| Cocaine Use Disorder, n/N (%) | 278/3696 (7.5) | 33/276 (12.0) | 20/154 (13.0) | 13/122 (10.7) |
| Alcohol Use Disorder, n/N (%) | 235/3696 (6.4) | 41/276 (14.9) | 24/154 (15.6) | 17/122 (13.9) |
| **Predicted Risk** |  |  |  |  |
| Pooled Cohort Equations 10-year risk score, Median (IQR) [N] | 6.6 (4.4, 11.1) [1681] | 9.0 (5.2, 14.5) [220] | 9.0 (5.2, 15.4) [122] | 9.0 (5.2, 13.1) [98] |
| Pooled Cohort Equations Lifetime risk score, Median (IQR) [N] | 45.5 (39.1, 50.4) [1204] | 39.1 (31.7, 50.4) [32] | 36.4 (26.9, 45.5) [19] | 50.4 (39.1, 50.4) [13] |
| Framingham risk score, Median (IQR) [N] | 12.1 (8.6, 20.6) [2266] | 15.8 (10.4, 25.6) [238] | 14.5 (10.1, 27.5) [130] | 16.4 (10.8, 24.2) [108] |
| **Baseline Values for Outcomes** |  |  |  |  |
| Systolic BP, mmHg, Median (IQR) [N] | 129 (120, 140) [3233] | 130 (118, 144) [247] | 127 (116, 144) [138] | 130.0 (120, 143) [109] |
| Diastolic BP, mmHg, Median (IQR) [N] | 80 (72, 85) [3233] | 79 (70, 88) [247] | 78 (69, 87) [138] | 79 (71, 89) [109] |
| LDL Cholesterol, Median (IQR) [N] | 98 (78, 123) [3235] | 101 (78, 128) [247] | 94.5 (74, 127) [138] | 104 (85, 132) [109] |

Footnotes: [1] Continuous variables are listed with Median (IQR). Number included is listed in brackets. Categorical variables are listed as frequency (percentage). [2] Baseline visit is defined as the first cardiology clinic visit or the last non-cardiology clinic appointment within each 6-month cohort. [3] Components are age, MI, CHF, PVD, DVA/TIA, Dementia, COPD, Connective tissue disease, Peptic ulcer disease, liver disease, diabetes, hemiplegia, moderate to severe chronic kidney disease,

### Lipid and Blood Pressure Control

At baseline, there were no differences in LDL (median 100 vs 97.5 mg/dL), but a higher proportion were prescribed lipid lowering therapy at baseline among those with a future cardiology clinic visit (22% vs 12%). The proportion prescribed antihypertensives increased to 24% after cardiology visit compared to 15% after the equivalent ambulatory visit. Cardiology clinic visits were not associated with subsequent lipid control over two years (55% vs 47%, adjusted OR 2.25, 95% CI 0.72-7.0, p=0.16, Figure 2), although with wide confidence intervals that do not exclude a possible benefit. Compared to those without a cardiology clinic visit, there was no significant difference in the change in LDL (3.7 mg/dl increase per year among those with a cardiology visit compared to 0.44 mg/dl increase per year among those without a cardiology encounter, or a between-group difference of 3.3 mg/dl per year, 95% CI −3 to 10, p=0.31, Table 2).

**Figure 2.**
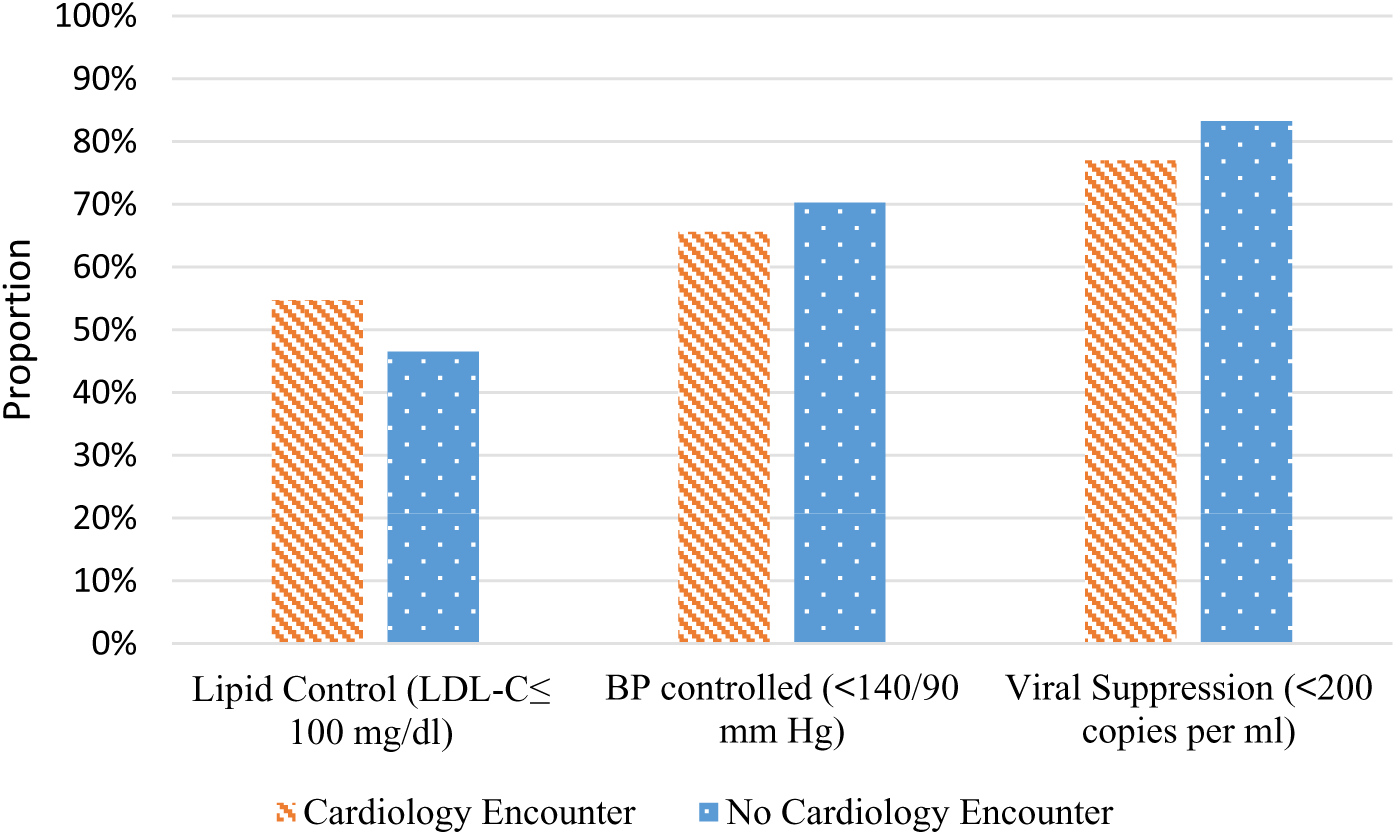
**Lipid, Blood Pressure, and HIV Control Over 2 Years by Cardiology Clinic Visit** Among UREG PWH with elevated cardiovascular risk, cardiology clinic visits were not associated with improved lipid control (adjusted OR 2.25, 95%CI 0.72-7.01, p= 0.16), blood pressure control (adjusted OR 0.78, 95% CI 0.49-1.24, p=0.29), or with the falsification endpoint of viral suppression, defined as either viral load <200 copies/ml or undetectable (adjusted OR 0.34, 95% CI 0.08-1.45, p=0.15).

**Table 2:** Change in Blood Pressure, LDL Cholesterol, and CD4 Count Per Year by Cardiology Clinic Visit. ^[1]^

| Outcome Measurement | Cardiology Visit | No Cardiology Visit | Adjusted Difference in Change per Year | P-value |
| --- | --- | --- | --- | --- |
| Systolic Blood Pressure, mm Hg per year<sup>[2]</sup> | 0.70 (-0.16, 1.55) | 0.21 (0.07, 0.34) | 0.49 (-0.38, 1.36) | 0.27 |
| Diastolic Blood Pressure, mm Hg per year<sup>[2]</sup> | 0.34 (-0.22, 0.90) | 0.31 (0.22, 0.40) | 0.03 (-0.53, 0.60) | 0.91 |
| LDL Cholesterol, mg/dL per year<sup>[3]</sup> | 3.70 (-2.53, 9.94) | 0.44 (-0.45, 1.33) | 3.26 (-3.03, 9.56) | 0.31 |
| **FALSIFICATION ENDPOINT** |  |  |  |  |
| CD4 Count, cells per mL per year<sup>[4]</sup> | 2.63 (-14.4, 19.7) | 14.4 (12.2, 16.6) | -11.8 (-28.9, 5.39) | 0.18 |

Footnotes: <sup>[1]</sup> Adjustment variables include age, sex, insurance (public, private, HRSA, other, no insurance), rural location, social deprivation index, HIV variables (CD4 count, viral suppression, anti-retroviral therapy), hepatitis C, Charlson Comorbidity Index, eGFR, BMI, diabetes, smoking, systolic blood pressure, and baseline LDL-C. Blood pressure analyses additionally adjust for anti-hypertensive treatment and lipid analyses additionally adjust for lipid lowering treatment at baseline. <sup>[2]</sup> N=3376 unique patients (244 with cardiology encounter). <sup>[3]</sup> N=2340 (159 with cardiology encounter) due to missing outcome assessments. <sup>[4]</sup> N=3100 unique patients included in this analysis, N=229 with cardiology visit.

At baseline, there were no differences in systolic blood pressure (median of 130 vs 129 mm Hg), but a higher proportion among those with a future cardiology clinic visit were prescribed antihypertensives at baseline (41% vs 28%). The proportion prescribed antihypertensives increased to 51% after cardiology visit compared to 29% after the equivalent ambulatory visit. Cardiology clinic visits were not associated with subsequent blood pressure control over two years (66% versus 70%, adjusted OR 0.78, 95% CI 0.49-1.24, p=0.29, Figure 2). Compared to those without a cardiology clinic visit, those with a cardiology clinic visit did not subsequently have lower increase in systolic blood pressure (0.7 mm Hg increase per year for patients with a cardiology visit, compared to 0.2 mm Hg increase per year for those without a cardiology visit, for a between-group difference of 0.5 mm Hg per year, 95% CI −0.4 to 1.4, p=0.27, Table 2).

**Figure 3.**
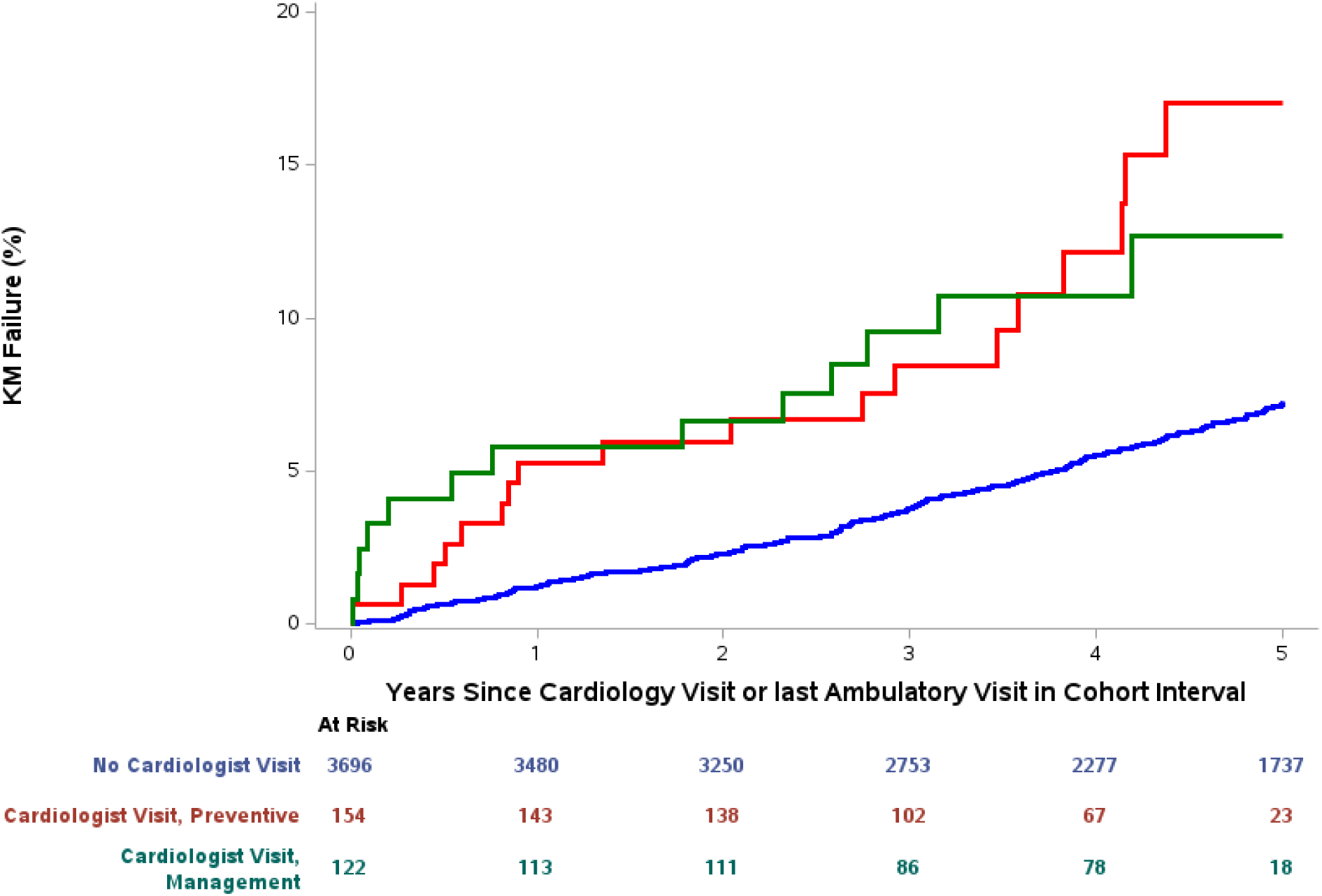
**Kaplan Meier Failure Curves for MACE by Cardiology Clinic Visit** Kaplan Meier plots of first MACE event by cardiology visit with the number at risk at each time interval below (No Cardiology Clinic Visit, Cardiology Clinic Visit-Prevention, and Cardiology Clinic Visit-Management). MACE events occurred earlier among those with a cardiology clinic visit; those with a management visit had earlier occurrence of first MACE, largely due to higher rates of coronary revascularization (see “Coronary Events” in Figure 4).

Results were similar for our falsification endpoints of viral suppression (Figure 2) and CD4 count (Table 2) with no difference in the proportion with viral suppression or in the CD4 count over two years among those with a cardiology clinic visit compared to those without a cardiology clinic visit.

**Table 3.**
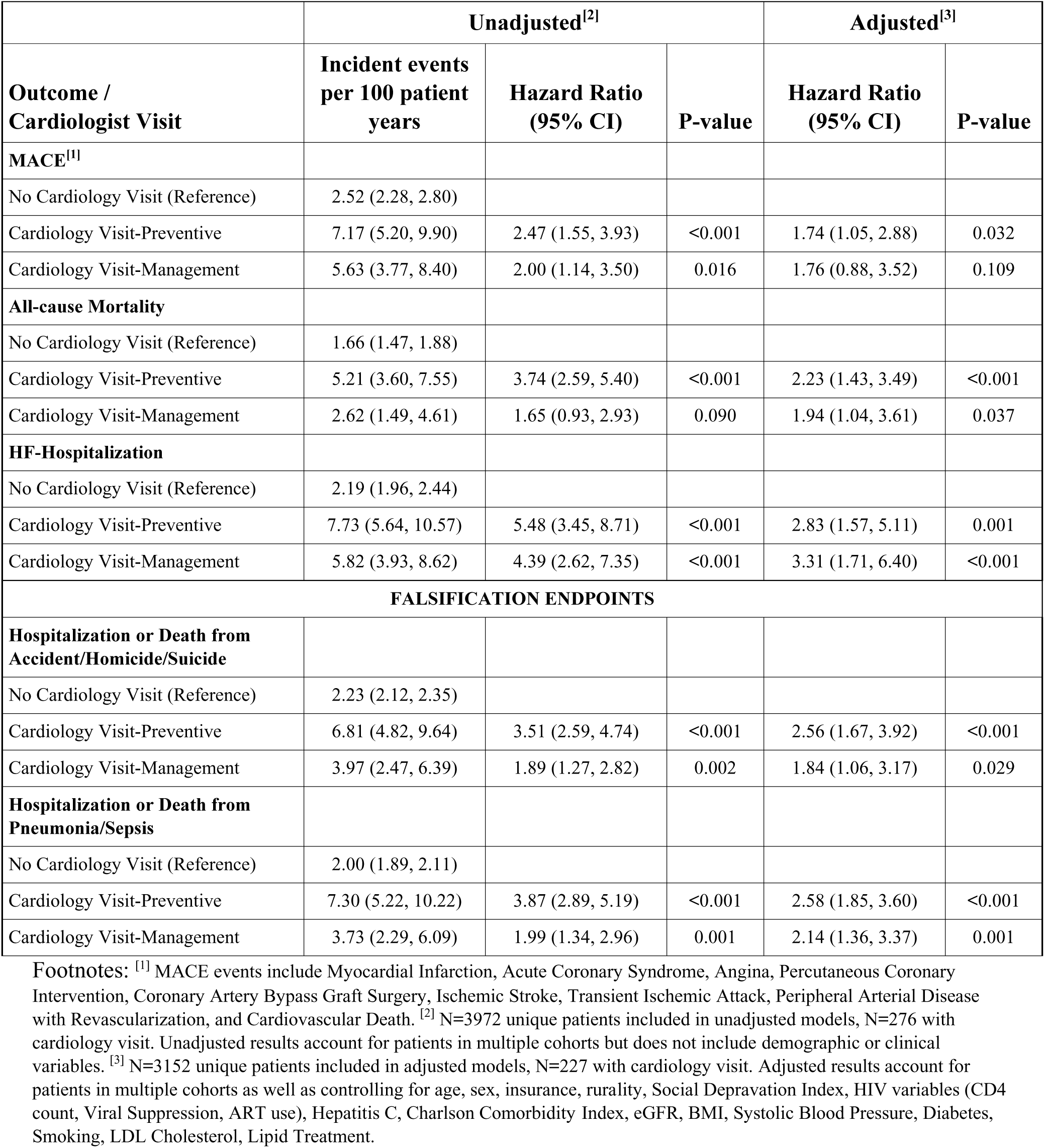
Incident Events by Cardiology Visit.

### Major Adverse Cardiovascular Events

Among 3,972 included patients, 276 (6.9%) had a cardiology clinic visit, of which 154 (56%) were classified as preventive and 122 (44%) were classified as management encounters. Over a median 5 years of follow-up, 237 individuals had a MACE event with 206 in the no cardiology clinic visit group and 31 in the cardiology clinic visit group. There were 2.5 incident MACE events per 100 patient years among those without a cardiology encounter (95% CI 2.3-2.8), 7.2 with a prevention cardiology encounter (95% CI 5.2-9.9), and 5.6 with a management cardiology encounter (95% CI 3.8-8.4). The unadjusted risk of MACE was 2.5 times higher among those with a prevention cardiology encounter (Hazard ratio (HR) 2.5, 95% CI 1.6-3.9) and 2.0 times higher for a management cardiology encounter (95% CI 1.1-3.5), than in patients without a cardiology visit. After adjustment for potential measured confounders, the hazard ratios were only slightly attenuated (adjusted HR 1.74, 95% CI 1.1-2.9 for prevention vs none and adjusted HR 1.76, 95% CI 0.9 to 3.5 for management vs none).

The risk of our falsification endpoints, which we hypothesized would not be impacted by a cardiologist clinic visit, were similarly higher among those with a cardiology clinic visit. The event rates per 100 patient years for hospitalization or death from accident, homicide, or suicide were 2.2 (95% CI 2.1-2.4), 6.8 (95% CI 4.8-9.6), and 4.0 (95% CI 2.5-6.4) for those without a cardiology clinic visit, those with a preventive cardiology clinic visit, and those with a management cardiology clinic visit, respectively. Compared to those without a cardiology clinic visit, the adjusted hazard ratios were 2.6 (95% CI 1.7-3.9) for preventive and 1.8 (95% CI 1.1-3.2) for management, compared to patients without a cardiology visit. The results for hospitalization or death from pneumonia or sepsis were nearly identical with event rates of 2.0 (95% CI 1.9-2.1), 7.3 (95% CI 5.2-10.2), and 3.7 (95% CI 2.3-6.1), and adjusted hazard ratios of 2.6 (95% CI 1.9-3.6) and 2.1 (95% CI 1.4-3.4), respectively. Results were similar in sensitivity analyses including non-cardiovascular mortality and heart failure in the MACE outcome (Supplemental Table 2).

### Type of Events by Cardiology Encounter

In a descriptive analysis, we examined the cumulative incidence of events within five years. Compared to those with a cardiology visit, the overall incident event rates were lower among those without a cardiology clinic visit with differences in the event types that occurred (Figure 4). Heart failure hospitalization had a similar cumulative incidence to non-cardiovascular mortality and MACE among those with a cardiology clinic visit, regardless of whether it was a management or prevention visit) but was less common among those without a cardiology clinic visit. Non-cardiovascular mortality was highest among those with a cardiology prevention clinic visit and revascularization was more common among those with a cardiology management clinic visit.

**Figure 4.**
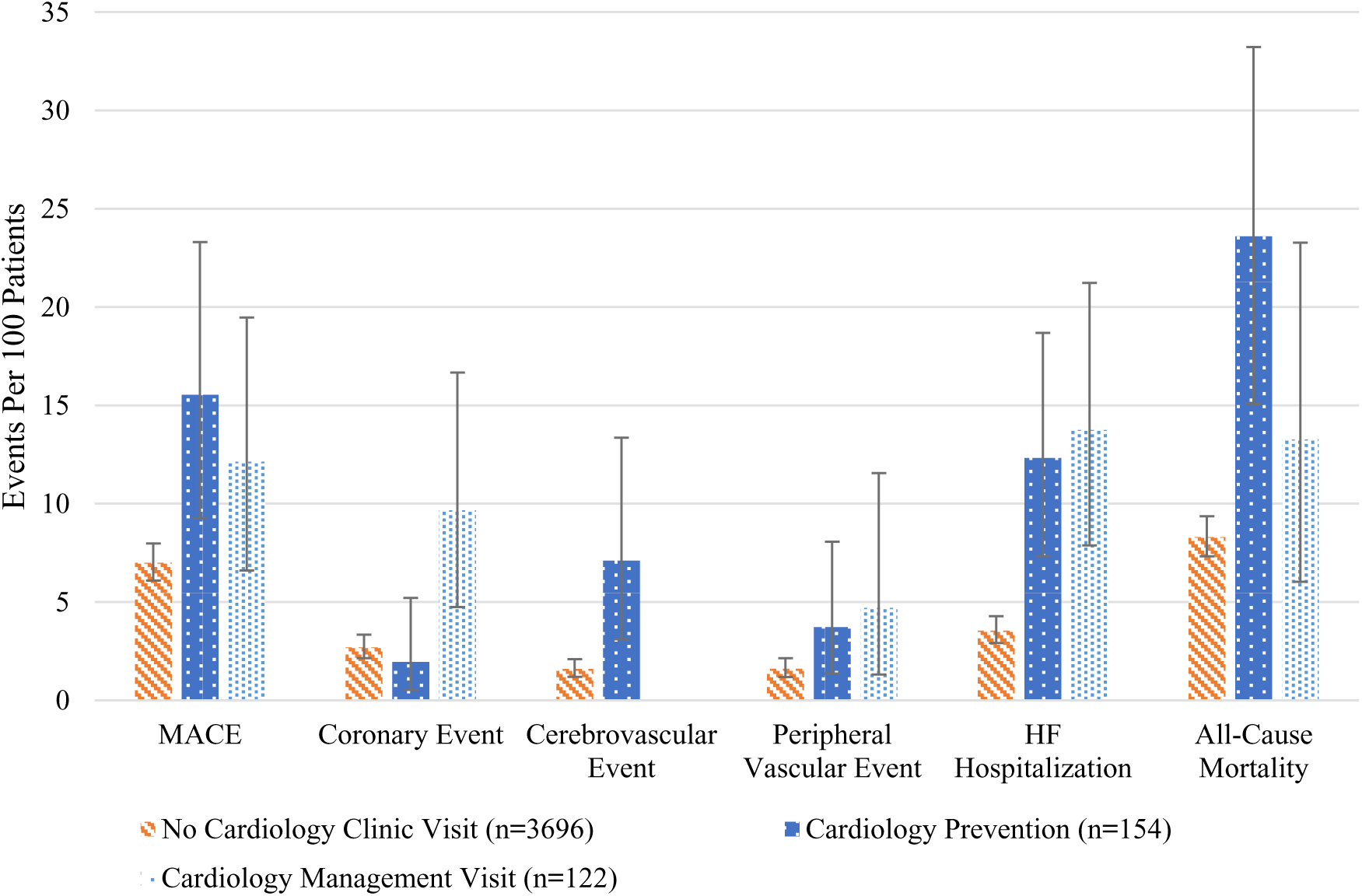
**Five Year Cumulative Incidence of Cardiovascular Events and Mortality By Cardiology Clinic Visit Group** Bar charts of cumulative incidence of MACE events, heart failure (HF) hospitalization, and all-cause mortality over 5 years of follow-up (bars represent events per 100 patient years and lines represent 95% CI). These events are not mutually exclusive; a single patient could have a coronary event, cerebrovascular event, and heart failure hospitalization within 5 years, for example. Coronary event included myocardial infarction, hospitalization for acute coronary syndrome, percutaneous coronary intervention, and coronary artery bypass graft surgery; cerebrovascular event included stroke and transient ischemic attack, and peripheral vascular event includes peripheral vascular disease requiring revascularization. All outcomes were more common among individuals with a cardiology encounter.

## Discussion

Among a cohort of PWH from UREGs without known cardiovascular disease across four health systems in the Southern United States, few patients had a “primary prevention” cardiology clinic visit prior to a major cardiovascular event or diagnosis; cardiology clinic visits were not associated with improvements in blood pressure or lipid control. Even accounting for higher baseline risk, those with a cardiology clinic visit had a higher subsequent risk of MACE, heart failure hospitalization, and mortality as well as our falsification endpoints than those without a cardiology clinic visit regardless of whether the first cardiology encounter was classified as a prevention or management visit.

To our knowledge, there are no prior studies establishing whether cardiology encounters result in better lipid control and blood pressure control (primordial prevention) or prevent future cardiovascular events (primary prevention). Compared to people without HIV, PWH have worse cardiovascular health as operationalized by the American Heart Association’s Life’s Essential 8.^9^ Earlier control of risk factors, or optimization of cardiovascular health, is associated with longer morbidity-free survival among PWH.^10^ Although the American Society for Preventive Cardiology has established clinical practice guidelines for primordial and primary prevention including the role of the preventive cardiologist to promote health equity,^11^ data are lacking on the association between cardiology clinic visits and cardiovascular outcomes among patients at risk for cardiovascular disease.

In this study, we found that a cardiologist encounter alone is not strongly associated with achievement of lipid control, defined as an LDL-C<100 mg/dL, or blood pressure control, defined as <140/90 mm Hg. Those with a cardiology clinic visit within our study had higher baseline rates of antihypertensive and lipid lowering prescriptions such that there were no differences in LDL or blood pressure at baseline. We expected that cardiologists would be more aggressive about prescribing lipid lowering therapy, and that statin prescriptions would be specific mechanism by which a single cardiology clinic visit could prevent cardiovascular events. Unexpectedly, the proportion prescribed lipid-lowering therapy was nearly identical after the cardiology encounter, while there was a modest increase in anti-hypertensive prescriptions, with no significant improvements in mean LDL or SBP or the proportion controlled, although the wide confidence intervals for LDL do not exclude a possible clinically significant benefit. Perhaps cardiologists were reluctant to prescribe statins due to concerns about drug-drug interactions with older antiretroviral therapy regimens, did not think statins were appropriate without clinical trial data to support their use among PWH, or did not feel that it was their role to prescribe them for primary prevention. Our qualitative research in the same population suggests that individual and structural stigma may impact the effectiveness of cardiology specialty care.^12^

It is important to note this study was conducted before completion of the Randomized Trial to Prevent Vascular Events in HIV (REPRIEVE), which demonstrated that pitavastatin reduced cardiovascular events among PWH at elevated cardiovascular compared to placebo,^13^ so it may be that cardiologists (as well as HIV and primary care physicians) would now be more aggressive about initiation of lipid lowering therapy. Guidelines have now shifted toward considering statins for all PWH over age 40 and recommending them for all with elevated cardiovascular risk.^3,14^

Our findings that cardiology clinic visits were associated with more than double the risk of MACE, even controlling for measured confounders, were unexpected. It is possible that referring providers are identifying and selectively referring those at higher risk of cardiovascular disease and overall mortality in ways that are not adequately captured in the EHR and thus could not be adequately accounted for in our adjusted models. A hypothetical patient vignette that may help explain how our findings diverged from our hypothesis is a patient with worsening chronic stable angina being managed by a primary care physician who subsequently sees a cardiologist and is referred for revascularization; such a patient would have a shorter time to a revascularization event in the cohort in which they saw a cardiologist. However, similar findings for all-cause mortality and most strikingly for our falsification endpoints suggest that the patients seen by cardiologists are at higher overall baseline risk. Another alternative explanation is that the cardiologist clinic visits could increase risk of MACE particularly via referral for procedures that have risk, but we would not expect the cardiology encounter would increase the risk of our falsification endpoints, so this explanation is less likely.

Our study highlights how there is equipoise regarding interventions and strategies to promote cardiovascular health and equity. It is challenging to adequately answer these causal questions with observational EHR-based research. One recent randomized clinical trial, A Nurse-Led Intervention to Extend the HIV Treatment Cascade for Cardiovascular Disease Prevention (EXTRA-CVD), of an implementation strategy for a nurse-led care coordination program that includes an electronic health record tools, home blood pressure monitoring, and an evidence-based treatment algorithm, improved blood pressure and lipid control over 12 months, something that we were not able to demonstrate in this observational study.^15^ Several features of EXTRA-CVD that may have contributed to its success were that the intervention was a multidisciplinary, nurse-led approach with longitudinal relationships aided by integrated EHR tools. Finally, by randomizing individuals EXTRA-CVD was able to ensure greater exchangeability between those who did and did not get the intervention to assess the causal impact of the intervention.

### Limitations

This is an observational comparative effectiveness study with limitations inherent to the study design. The use of EHRs is a reasonable approach to study the question of a whether cardiology clinic visits (which are easily measurable in the EHR) are associated with process and outcome measures, including blood pressure and lipids which are often measured within the EHR, but it still comes with serious drawbacks. For example, capturing the factors that lead to cardiology referral without access to the clinician and patient interaction (or even the documentation of the referral in the EHR) makes the risk of confounding by indication challenging to overcome, and even our qualitative work in this patient population only captures the perspectives of those who completed their cardiology referral.^12^ Nonetheless, we tried to stratify our exposure variable by the diagnoses coded by the cardiologist, which may have been misclassified and depends on the coding strategy of the cardiologists and was likely subject to misclassification. We did not have data on referral to cardiology to identify time from referral to cardiology clinic visit, to classify those who were referred but did not see a cardiologist as exposed, or to identify those who had MACE events or died while waiting to see a cardiologist. In our previous work in this cohort, we observed a long delay between eligibility (onset of elevated cardiovascular risk) and cardiology encounter (~2 years)^4^, so we had to develop an analytic strategy to prevent immortal time bias in our outcomes analysis. Our use of a sequential cohort design and using equivalent non-cardiology ambulatory encounters partially addresses the issues of immortal time bias but does not address the concern of those who had MACE events occurring between cardiology referral, which was unmeasured, and scheduled cardiology clinic visit. Because the factors leading to cardiology encounters were not well-captured, we could not accurately develop a model to predict propensity for cardiology encounters for a matching-based strategy to maximize exchangeability between groups. Because our study population is limited to PWH who identify as UREGs who live in the South, these findings may not be externally generalizable to all PWH, people without HIV, or people in other regions of the country where discrimination or other structural factors attenuate the efficacy of a cardiology clinic visit in ways specific to the included patient population.

### Conclusions

In conclusion, our study demonstrates that for PWH who identify as members of UREGs, a single cardiology clinic visit is not strongly associated with improved lipid or blood pressure control. Those who have a cardiology clinic visit have higher risk of adverse outcomes including MACE, mortality, and non-cardiovascular outcomes. Whether prospective referral of PWH at elevated cardiovascular risk would result in optimization of cardiovascular health and prevention of cardiovascular events cannot be determined from our observational study and is an important area for further research, but our findings suggest that a single cardiology encounter may not have a large effect on cardiovascular health in this population.

## Data Availability

A de-identified dataset is currently being generated to be posted to the NIH's BioData Catalyst.

## Acknowledgements

None.

## Presentation

Preliminary results were presented at the American Heart Association Epidemiology and Lifestyle Scientific Sessions in March 2024 in Chicago, IL.

